# Real-world Effectiveness of 2-dose SARS-CoV-2 Vaccination in Kidney Transplant Recipients

**DOI:** 10.1101/2021.09.21.21263457

**Authors:** Caitríona M. McEvoy, Anna Lee, Paraish S. Misra, Gerald Lebovic, Ron Wald, Darren A. Yuen

## Abstract

The humoral response to two doses of SARS-CoV-2 (Covid-19) vaccine among transplant recipients is inferior to immunocompetent individuals.^1^ Data on the ‘real-world’ effectiveness of vaccination in kidney transplant recipients [KTRs] are lacking. We performed a cohort study to investigate the impact of vaccination on Covid-19 infection and outcomes in our kidney transplant program.

## Main Text

Covid-19 vaccination and infection status between March 11, 2020 and July 19, 2021 were recorded for all KTRs in our program (Supplemental Fig.1), along with demographics and clinical covariates associated with Covid-19 severity. A time-varying Cox-Proportional Hazards model examined the association between vaccine status and the risk of Covid-19 infection, adjusting for age, sex, time post-transplant, kidney function, Covid-predisposing comorbidities, and the weekly infection rate in each patient’s community. The secondary outcome was severe Covid-19 infection, defined as Covid-related hospital admission or death. Detailed methods are described in the Supplementary Appendix.

We evaluated 1793 KTRs. 1540 (85.9%) KTRs had received 1 vaccine dose, and 1402(78.2%) had received 2 doses by study end. The median age was 60.4 years (interquartile range [IQR], 51.0 to 69.2), and the median time from transplantation was 8.1 years (IQR, 3.9 to 13.6). Supplementary Table 1 summarizes KTR baseline characteristics.

There were 114 Covid-19 infections, of which 61% were severe. Community infection rates (Hazard Ratio [HR] 1.08 per 10 in 100,000 increase in weekly infection rate; 95% confidence interval [CI], 1.04 to 1.12) and transplant duration <3 months (HR 4.29; CI,1.5 to 12.23) were associated with infection risk (Table 1, Supplemental Fig.4). Similar associations were observed for severe Covid infections (Supplemental Table 6). As compared to unvaccinated patients, we did not detect an association between double vaccination and any Covid-19 infection (HR 0.52 (95%CI, 0.14 to 1.95) or severe infection (HR 0.59 (95%CI, 0.12 to 2.89). A sensitivity analysis examined the effect of administration of any vaccine dose on any Covid-19 infection (HR 0.78; 95%CI, 0.38 to 1.61; p=0.5) and severe infection (HR 1.22; 95%CI,0.53 to 2.8; p=0.64), and found no change in overall result.

**Table 1:** Association between vaccine status and the risk of contracting Covid-19.

| Variable | HR | Lower CI | Upper CI | p-value |
| --- | --- | --- | --- | --- |
| Vaccine dose 1 | 0.87 | 0.41 | 1.87 | 0.73 |
| Vaccine dose 2 | 0.60 | 0.15 | 2.31 | 0.45 |
| Age | 0.99 | 0.98 | 1.01 | 0.36 |
| Transplant duration >3 months | 1.24 | 0.51 | 2.30 | 0.64 |
| Transplant duration 0-3 months | 4.29 | 1.50 | 12.23 | 0.007 |
| Female sex | 1.19 | 0.81 | 1.73 | 0.36 |
| Latest eGFR | 0.99 | 0.98 | 1.00 | 0.11 |
| CDC-score | 1.03 | 0.89 | 1.19 | 0.73 |
| Covid community infection burden | 1.08 | 1.04 | 1.12 | <0.001 |

Although our analysis of vaccination effectiveness may be limited by cohort size, our results clearly show that the risks of infection and poor outcome were predominantly determined by community infection rates and transplant vintage, rather than by vaccination status. Recent work demonstrates that a third vaccine dose yields greater immunogenicity in transplanted individuals^2-5^. Future studies must examine whether such strategies enhance real-world vaccine effectiveness among transplant patients.

Association between vaccine status and the risk of contracting Covid-19 after adjusting for age (per one year increase), sex, transplant duration, latest eGFR and geographical Covid-19 burden. The following were treated as time varying variables: Vaccine (none, one, two doses); Transplant duration (0-3 months, 3-12 months, >12 months); and Covid-19 community infection burden per 100,000 population (HR reflects an increase in 10 per 100,000 population).

## Data Availability

Data regarding Covid-19 rates in Ontario is publicly available at (https://www.publichealthontario.ca/)

## Abbreviations

CDC: Centers for Disease Control
CI: Confidence Interval
eGFR: estimated Glomerular Filtration Rate
HR: Hazard Ratio

## References

1. Boyarsky, B.J., et al. Antibody Response to 2-Dose SARS-CoV-2 mRNA Vaccine Series in Solid Organ Transplant Recipients. JAMA 325, 2204–2206 (2021).

2. Hall, V.G., et al. Humoral and cellular immune response and safety of two-dose SARS-CoV-2 mRNA-1273 vaccine in solid organ transplant recipients. Am J Transplant (2021).

3. Werbel, W.A., et al. Safety and Immunogenicity of a Third Dose of SARS-CoV-2 Vaccine in Solid Organ Transplant Recipients: A Case Series. Ann Intern Med (2021).

4. Kamar, N., et al. Three Doses of an mRNA Covid-19 Vaccine in Solid-Organ Transplant Recipients. N Engl J Med 385, 661–662 (2021).

5. Del Bello, A., et al. Efficiency of a boost with a third dose of anti-SARS-CoV-2 messenger RNA-based vaccines in solid organ transplant recipients. Am J Transplant (2021).

