## Supplementary Appendix for "Real-world Effectiveness of 2-dose SARS-CoV-2 Vaccination in Kidney Transplant Recipients"

**Table of Contents**

**Methods**

Patient population and study design

Data sources

Outcomes

Statistical analysis

**Supplementary Tables (ST)**

ST1: Demographic characteristics and clinical features of the study population

ST2: Outcomes in COVID-19-infected patients

ST3: Causes of ESKD in study population

ST4: Burden of CDC-defined severe-covid predisposing conditions in study population

ST5: Breakdown of vaccination status in KTR cohort.

ST6: Association between vaccination and the risk of severe Covid-19 infection

ST7: Sensitivity analysis examining association between any vaccine dose and the risk of Covid-19 infection

ST8: Sensitivity analysis examining association between any vaccine dose and the risk of severe Covid-19 infection

**Supplementary Figures (SF)**

SF1: Study flow chart

SF2: Comparison of Covid-19 cases in Ontario, and in kidney transplant recipients

SF3: Graph of Covid-19 strains over time

SF4: Probability of Covid-19 infection over time stratified by neighbourhood infection rate and transplant duration.

**Appendices**

Appendix 1: Email Survey sent to all kidney transplant recipients

**References**

**Methods**

**Patient population and study design**

We conducted a retrospective cohort study of kidney transplant recipients followed by the Renal Transplant Program of St. Michael’s Hospital (Unity Health Toronto), Toronto, Ontario. This initiative was formally reviewed by institutional authorities at Unity Health Toronto and deemed to require neither Research Ethics Board approval nor written informed consent from participants.

We surveyed adult patients (aged ≥ 18 years) who had received a kidney transplant and had a functioning allograft. All such patients were considered eligible for inclusion. Our survey encompassed all prevalent patients as of June 7, 2021, and incident patients who received transplants between June 7, 2021 and July 19, 2021.

Patients were contacted by telephone or email, and surveyed (Appendix 1) to determine the details of their Covid-19 vaccination and infection status, and outcomes of Covid-19 infection between the period of March 11, 2020 to July 19, 2021. The date of March 11, 2020 was chosen as this was the date that the World Health Organization declared worldwide Covid-19 infections as a global pandemic. The email and/or phone surveys were conducted between June 7, 2021 and July 19, 2021.

**Data sources**

Patient demographics, clinical characteristics and covariates, and any missing, conflicting or incomplete data related to Covid-19 vaccination or infection status were collected from the transplant patient clinical database and electronic health records.

Follow-up for each patient started from the patient’s transplant date or Mar 11, 2020, whichever was later.

Transplant duration was calculated as the time from the date of transplant, to the earlier of 1) Date of Covid-19 infection, or 2) July 19^th^ 2021.

The initial cause for end stage kidney disease for each patient was assigned to one or more of 6 broad categories (Glomerulonephritis, Diabetic Kidney Disease, Hypertension, Polycystic Kidney Disease, Thrombotic Microangiopathy, and Other).

A record of the weekly community rates of infection for each of Ontario’s 34 public health units is publicly available from (<https://www.publichealthontario.ca/en/data-and-analysis/infectious-disease/covid-19-data-surveillance/covid-19-data-tool?tab=trends>). These data were used to compile quartiles of community infection risk, as depicted in Supplemental Fig. 4A.

Patients were assigned to their respective community (public-health unit) by the postal-code of their place of residence, using the online tool available at (<https://www.phdapps.health.gov.on.ca/phulocator/>).

The co-morbidities likely to influence Covid-19 infection and severity were obtained from the list of conditions identified by the Centers for Disease Control and Prevention (CDC) obtained from (<https://www.cdc.gov/coronavirus/2019-ncov/need-extra-precautions/people-with-medical-conditions.html>). This list includes 17 broad categories of diseases. All patients in our Renal Transplant Clinic automatically fulfilled criteria for three categories including Chronic Kidney Disease, Immunocompromised State, and Solid Organ or Blood Stem Cell Transplant. We calculated a score (‘CDC-score’) for the total number of coexisting conditions identified by the CDC as risk factors for severe Covid-19.

Information on Covid-19 strain was available for a subset of patients only. These data were incomplete mainly because testing for variants of concern was not widespread in Ontario prior to January 2021.

**Outcomes**

The primary outcome was Covid-19 infection in our study population. Severe Covid-19 infection (as defined by COVID-19 infection complicated by hospitalization and death) was analysed as a secondary outcome.

**Statistical Analysis**

Population demographics and clinical covariates were summarized using descriptive statistics.

A time-varying Cox Proportional Hazards (Cox-PH) model examined the relationship between vaccine status and the primary outcome of contracting Covid-19. Vaccine status was categorized in 3 levels: no dose (zero doses or <14 days following first dose), one dose (>14 days after their first dose, and/or <14 days after their second dose), and two doses (>14 days after their second dose), and was treated as a time-dependent variable. The model was parametrized to interpret the additional effect of a second dose as compared to a single dose. A contrast was used to estimate the effect of two doses as compared to zero doses of vaccine. Transplant vintage, coded as less than three months, three to twelve months and greater than 12 months; as well as the weekly incidence of Covid-19 were treated as time-dependent variables. The model also adjusted for age, sex, most recent eGFR value and CDC score. Schoenfeld residuals were used to test the proportional hazards assumption in the time invariant variables, and the proportional hazards assumption was met. A similar analysis was conducted examining the association of vaccine status and severe Covid-19 infection, defined as infection with hospitalization or death. A sensitivity analysis was performed treating vaccine status as a binary variable comparing at least one dose versus no doses.

Complete case analysis was used. Three participants died before contracting Covid-19. They were censored at their death time and included in the Cox Proportional hazards model, as the model is valid in the context of competing risk.^1^ Survival curves were created for Covid-19 community infection burden and transplant vintage, and were computed by exponentiating the hazard function.

Analyses were done in the R statistical environment (Version 4.0.2).^2^

**Supplementary Tables**

**Supplementary Table 1. Demographic characteristics and clinical features of the study population**

| Characteristic | Total study population  N=1793 | Covid-19  Infected  N=114 | Non-infected  N=1679 |
| --- | --- | --- | --- |
| Male sex, n (%) | 1141 (63.5%) | 71 (59.7%) | 1070 (63.7%) |
| Group/residential living | 9 (0.5%) | 5 (4.4%) | 4 (0.2%) |
| Transplant Type |  |  |  |
| Living donor, n (%) | 764 (42.7%) | 34 (29.8%) | 730 (43.5%) |
| Deceased donor, n (%) | 1029 (57.3%) | 80 (70.2%) | 949 (56.5%) |
| Latest eGFR (ml/min/1.73m^2^) | 54.1 (39.7, 70.1) | 51.7 (36.8,69.1) | 54.4 (40.0, 70.4) |
| Number of CDC-identified medical conditions,  median (IQR) | 5 (5, 6) | 5 (5, 6) | 5 (5, 6) |
| Age (years), median (IQR) | 60.4 (51.0, 69.2) | 60.0 (47.7, 69.3)^§^ | 60.4 (51.0, 69.2) |
| Transplant duration (years), median (IQR) | 8.1 (3.9, 13.6) | 7.3 (2.5, 12.7)^§^ | 8.1 (3.9, 13.7) |
| Covid-19 Vaccination Status |  |  |  |
| 1st Dose Vaccinated | 1543 (85.8%) | 26 (22.8%)^§^ | 1459 (86.9%) |
| 2^nd^ Dose Vaccinated | 1404 (78.1%) | 5 (4.4%)^§^ | 1336 (79.6%) |

§- These values are at the time of Covid-19 infection. The remaining values are calculated at study close.

Abbreviations: CDC, Centers for Disease Control; Covid-19, SARS-CoV-2; eGFR, Estimated Glomerular Filtration Rate; IQR, Inter-quartile range.

**Supplementary Table 2. Outcomes: Hospitalizations, Intensive Care Unit (ICU) Admissions, Intubation Status, and Deaths by Vaccination Status in 114 Covid-19-infected Patients**

| Vaccination Status | Infected | Hospitalized | ICU | Intubation | Death |
| --- | --- | --- | --- | --- | --- |
| No doses | 88 | 50 | 23 | 14 | 16 |
| <= 14 days post 1st dose | 12 | 8 | 2 | 2 | 2 |
| >14 days post 1st dose, no 2nd dose | 9 | 8 | 1 | 1 | 0 |
| <= 14 days post-2nd dose | 2 | 2 | 1 | 1 | 1 |
| 15-21 days post 2nd dose | 1 | 1 | 1 | 1 | 1 |
| >21 days post-2nd dose | 2 | 1 | 0 | 0 | 0 |
| Total | 114 | 70 | 28 | 19 | 20 |

Abbreviations: ICU, Intensive Care Unit

**Supplementary Table 3: Causes of ESRD in Study Population**

| Cause of ESRD in study population* | N |
| --- | --- |
| Glomerulonephritis | 764 |
| Diabetic Kidney Disease | 295 |
| Hypertension | 201 |
| Polycystic Kidney Disease | 193 |
| Thrombotic Microangiopathy | 8 |
| Other | 382 |

*Patients may be entered in >1 category if dual pathology present

Abbreviations: ESRD, End-Stage Renal Disease

**Supplementary** **Table 4**: **Distribution of CDC-identified medical conditions associated with increased severity of Covid-19 illness**

| CDC-identified Medical Condition | Infected  (n=114) | Non-Infected  (n=1679) |
| --- | --- | --- |
| Cancer, n(%) | 11 (9.7) | 254 (15.1) |
| Chronic Kidney Disease, n(%) | 114 (100) | 1679 (100) |
| Chronic Lung diseases, including COPD, asthma (moderate-to-severe), interstitial lung disease, cystic fibrosis, and pulmonary hypertension, n(%) | 7 (6.1) | 109 (6.5) |
| Dementia or other neurological conditions, n(%) | 8 (7) | 113 (6.7) |
| Diabetes, n(%) | 58 (50.8) | 693 (41.3) |
| T1DM, n(%) | 0 (0) | 28 (1.7) |
| T2DM, n(%) | 43 (37.7) | 490 (29.2) |
| DM- Not otherwise specified, n(%) | 15 (13.2) | 175 (10.4) |
| Down Syndrome, n(%) | 0 (0) | 0 (0) |
| Heart conditions (such as heart failure, coronary artery disease, cardiomyopathies, or hypertension), n(%) | 80 (70.2) | 1136 (67.7) |
| HIV Infection, n(%) | 0 (0) | 13 (0.8) |
| Immunocompromised State, n(%) | 114 (100) | 1679 (100) |
| Liver Disease, n(%) | 8 (7) | 92 (5.5) |
| Overweight (BMI 25-30), Obesity (30-40), Severe Obesity (BMI >40), n(%) | 57 (50) | 906 (54) |
| Overweight (BMI 25-30), n(%) | 32 (28.1) | 562 (33.5) |
| Obesity (BMI 30-40), n(%) | 20 (17.5) | 314 (18.7) |
| Severe Obesity (BMI>40) n(%) | 5 (4.4) | 30 (1.8) |
| Pregnancy, n(%) | 0 (0) | 4 (0.2) |
| Sickle cell disease or thalassemia, n(%) | 5 (4.4) | 32 (1.9) |
| Smoking, current, or former, n(%) | 30 (26.3) | 553 (32.9) |
| Solid Organ or Blood Stem Cell Transplant, n(%) | 114 (100) | 1679 (100) |
| Stroke or cerebrovascular disease, which affects blood flow to the brain, n(%) | 11(9.7) | 113 (6.7) |
| Substance use disorders, n(%) | 1 (0.9) | 13 (0.77) |

Abbreviations: BMI, Body Mass Index; CDC, Centers for Disease Control; COPD, Chronic Obstructive Pulmonary Disease; T1DM/T2DM; Type 1 and Type 2 Diabetes Mellitus, respectively.

**Supplementary Table 5: Covid-19 Vaccination Status at the time of survey administration**

|  | **N (%)** |
| --- | --- |
| **1st Dose Vaccinated** | 1540 (85.9%) |
| Pfizer-BioNTech Covid-19 mRNA vaccine (Tozinameran or BNT162b2) | 1184 |
| Moderna Covid-19 vaccine (mRNA-1273) | 275 |
| AstraZeneca Covid-19 vaccine (ChAdOx1-S) | 76 |
| Other | 5 |
| **2^nd^ Dose Vaccinated** | 1402 (78.2%) |
| Pfizer-BioNTech Covid-19 mRNA vaccine (Tozinameran or BNT162b2) | 1060 |
| Moderna Covid-19 vaccine (mRNA-1273) | 289 |
| AstraZeneca Covid-19 vaccine (ChAdOx1-S) | 53 |
| **Among Patients Vaccinated with 2 Doses** | 1402 |
| **Both** Pfizer-BioNTech Covid mRNA vaccine (Tozinameran or BNT162b2) | 1048 |
| **First dose:** Pfizer-BioNTech Covid-19 mRNA vaccine (Tozinameran or BNT162b2);  **Second dose:** Moderna Covid-19 vaccine (mRNA-1273) | 29 |
| **First dose:** Pfizer-BioNTech Covid-19 mRNA vaccine (Tozinameran or BNT162b2);  **Second dose:** AstraZeneca Covid-19 vaccine (ChAdOx1-S) | 1 |
| **Both** Moderna Covid-19 vaccine (mRNA-1273) | 253 |
| **First dose:** Moderna Covid-19 vaccine (mRNA-1273)  **Second dose:** Pfizer-BioNTech Covid-19 mRNA vaccine (Tozinameran or BNT162b2) | 5 |
| **Both** AstraZeneca Covid-19 vaccine (ChAdOx1-S) | 48 |
| **First dose:** AstraZeneca Covid-19 vaccine (ChAdOx1-S)  **Second dose:** Pfizer-BioNTech Covid-19 mRNA vaccine (Tozinameran or BNT162b2) | 7 |
| **First dose:** AstraZeneca Covid-19 vaccine (ChAdOx1-S)  **Second dose:** Moderna Covid-19 vaccine (mRNA-1273) | 7 |
| **First dose:** Other  **Second dose:** AstraZeneca Covid-19 vaccine (ChAdOx1-S) | 4 |

**Supplementary Table 6: Association between vaccine status and the risk of severe Covid-19 infection (i.e. Covid-19 infection associated with hospitalization or death)**

| **Variable** | **HR** | **Lower CI** | **Upper CI** | **p-value** |
| --- | --- | --- | --- | --- |
| Dose 1 | 1.48 | 0.62 | 3.52 | 0.38 |
| Dose 2 | 0.40 | 0.08 | 1.97 | 0.26 |
| Age | 1.01 | 0.99 | 1.03 | 0.25 |
| Transplant duration >3 months | 1.30 | 0.42 | 4.01 | 0.65 |
| Transplant duration 0-3 months | 6.19 | 1.87 | 20.49 | 0.003 |
| Female sex | 1.09 | 0.67 | 1.77 | 0.74 |
| Latest eGFR | 0.99 | 0.98 | 1.00 | 0.12 |
| CDC-score | 1.06 | 0.88 | 1.27 | 0.56 |
| Covid community infection burden | 1.10 | 1.05 | 1.15 | <0.001 |

| **Contrast** | **HR** | **Lower CI** | **Upper CI** | **p-value** |
| --- | --- | --- | --- | --- |
| Double vaccine Vs. no vaccine | 0.59 | 0.12 | 2.90 | 0.51 |

Association between vaccine status and the risk of severe Covid-19 infection (as defined by Covid-19 infection associated with hospitalization, death) after adjusting for age (per one year increase), sex, transplant duration, latest eGFR and geographical Covid burden. The following were treated as time varying variables: Vaccine (none, one, two doses); Transplant duration (0-3 months, 3-12 months, >12 months); and Covid community infection burden per 100,000 population (HR reflects an increase in 10 per 100,000 population).

Abbreviations: CDC, Centers for Disease Control; CI, Confidence Interval; eGFR, estimated Glomerular Filtration Rate

**Supplementary Table 7: Sensitivity analysis examining association between any vaccine dose and the risk of Covid-19 infection**

| **Variable** | **HR** | **Lower CI** | **Upper CI** | **p-value** |
| --- | --- | --- | --- | --- |
| Any vaccine dose | 0.78 | 0.38 | 1.61 | 0.50 |
| Age | 0.99 | 0.98 | 1.01 | 0.37 |
| Transplant duration >3 months | 1.23 | 0.51 | 2.97 | 0.65 |
| Transplant duration 0-3 months | 4.19 | 1.47 | 11.99 | 0.007 |
| Female sex | 1.19 | 0.81 | 1.73 | 0.38 |
| Latest eGFR | 0.99 | 0.98 | 1.00 | 0.11 |
| CDC-score | 1.03 | 0.89 | 1.19 | 0.73 |
| Covid community infection burden | 1.01 | 1.00 | 1.01 | <0.001 |

Sensitivity analysis examining association between receipt of any vaccine dose and the risk of contracting Covid-19 after adjusting age (per one year increase), sex, transplant duration, latest eGFR and geographical Covid-19 burden. The following were treated as time varying variables: Vaccine (none, one, two doses); Transplant duration (0-3 months, 3-12 months, >12 months); and Covid-19 community infection burden per 100,000 population (HR reflects an increase in 10 per 100,000 population).

Abbreviations: CDC, Centers for Disease Control; CI, Confidence Interval; eGFR, estimated Glomerular Filtration Rate

**Supplementary Table 8: Sensitivity analysis examining association between any vaccine dose and the risk of severe Covid-19 infection**

| **Variable** | **HR** | **Lower CI** | **Upper CI** | **p-value** |
| --- | --- | --- | --- | --- |
| Any vaccine dose | 1.22 | 0.53 | 2.80 | 0.64 |
| Age | 1.01 | 0.99 | 1.03 | 0.24 |
| Transplant duration >3 months | 1.3 | 0.42 | 3.99 | 0.66 |
| Transplant duration 0-3 months | 5.88 | 1.77 | 19.58 | 0.00 |
| Female sex | 1.09 | 0.67 | 1.76 | 0.74 |
| Latest eGFR | 0.99 | 0.98 | 1.00 | 0.13 |
| CDC-score | 1.06 | 0.88 | 1.27 | 0.57 |
| Covid community infection burden | 1.009 | 1.005 | 1.014 | <0.001 |

Sensitivity analysis examining association between receipt of any vaccine dose and the risk of poor outcome (hospitalization, death) following Covid-19 after adjusting age (per one year increase), sex, transplant duration, latest eGFR and geographical Covid-19 burden. The following were treated as time varying variables: Vaccine (none, one, two doses); Transplant duration (0-3 months, 3-12 months, >12 months); and Covid-19 community infection burden per 100,000 population (HR reflects an increase in 10 per 100,000 population).

Abbreviations: CDC, Centers for Disease Control; CI, Confidence Interval; eGFR, estimated Glomerular Filtration Rate

**Supplementary Figures**

**SF1: Study flow chart**

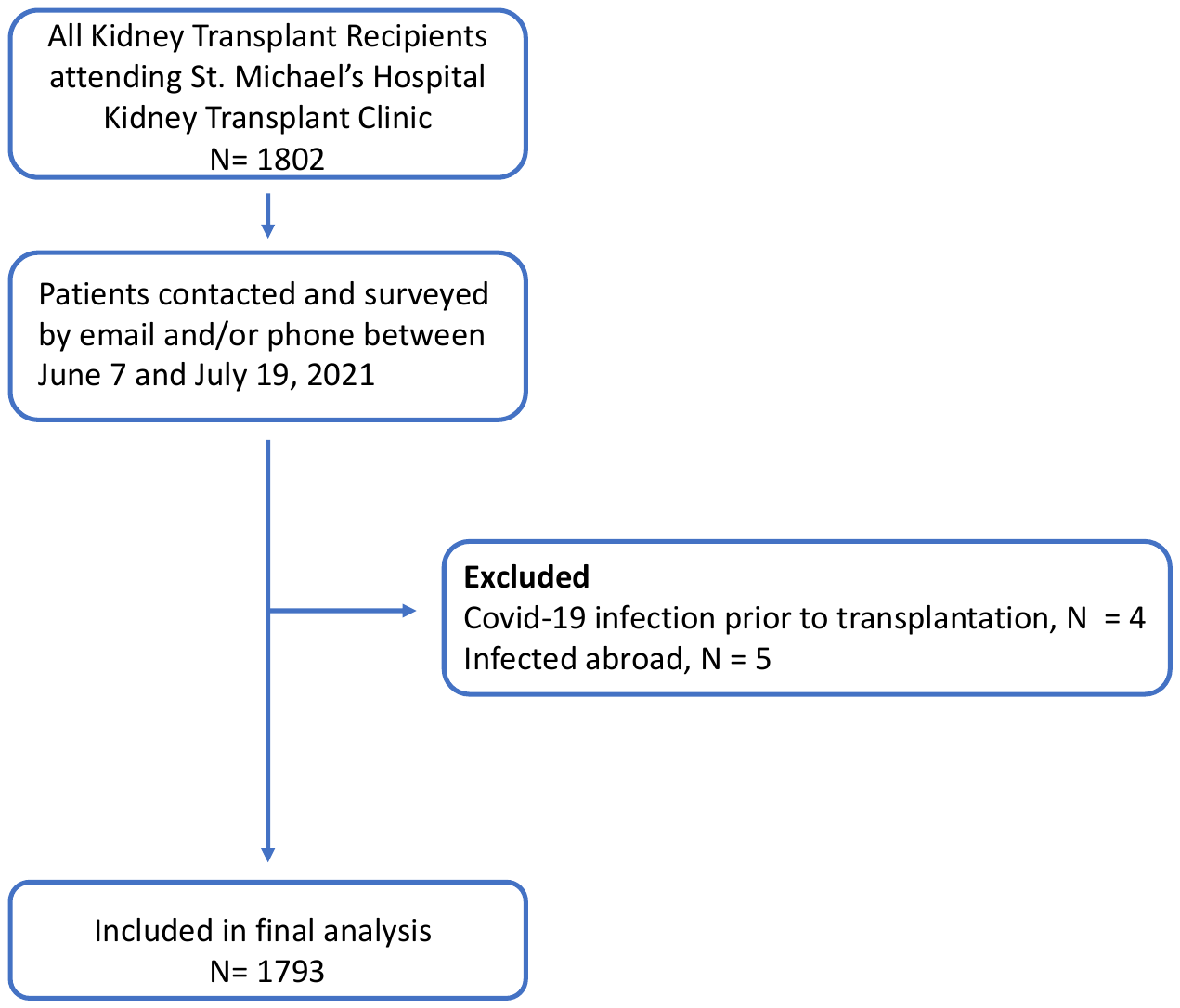

**SF2: Comparison of Covid-19 cases in Ontario, and in kidney transplant recipients throughout the study period.**

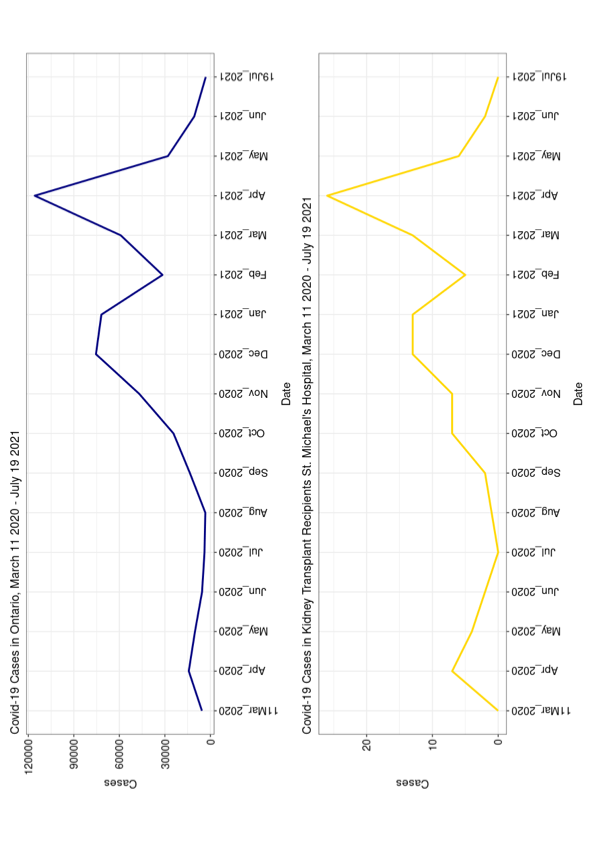

Comparison of Covid-19 cases in Ontario (top, in navy), and in kidney transplant recipients attending St. Michael’s Renal Transplant Clinic (bottom, in yellow) during the study period (March 11, 2020 – July 19, 2021). Ontario case data is publicly available from (<https://www.publichealthontario.ca>).

**SF3: Graph of Covid-19 strains over time**

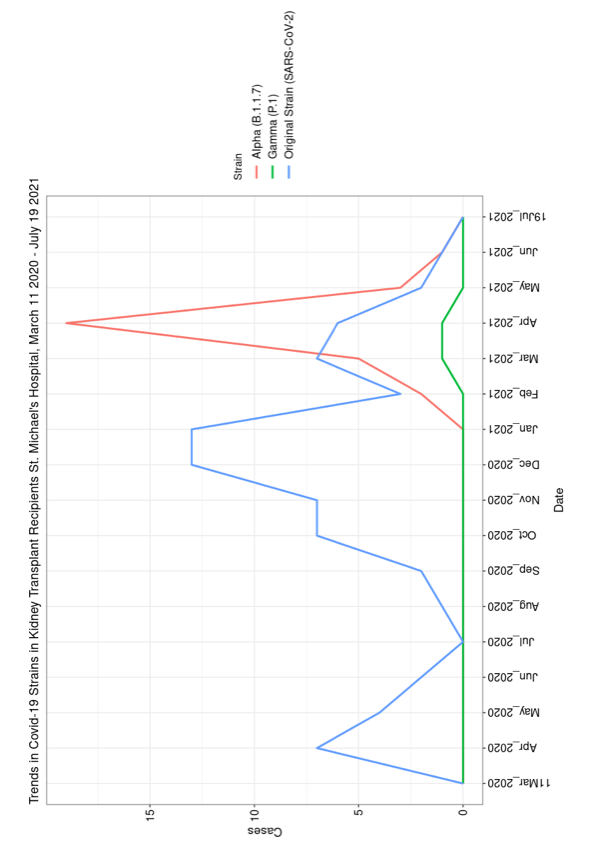

Variant testing was available for a subset of infected patients, and graphed over time. The majority of tested cases were the original Covid-19 strain (SARS-CoV-2) or the Alpha strain (B.1.1.7).

**SF4: Probability of Covid-19 infection over time stratified by neighbourhood infection rate and transplant duration.**

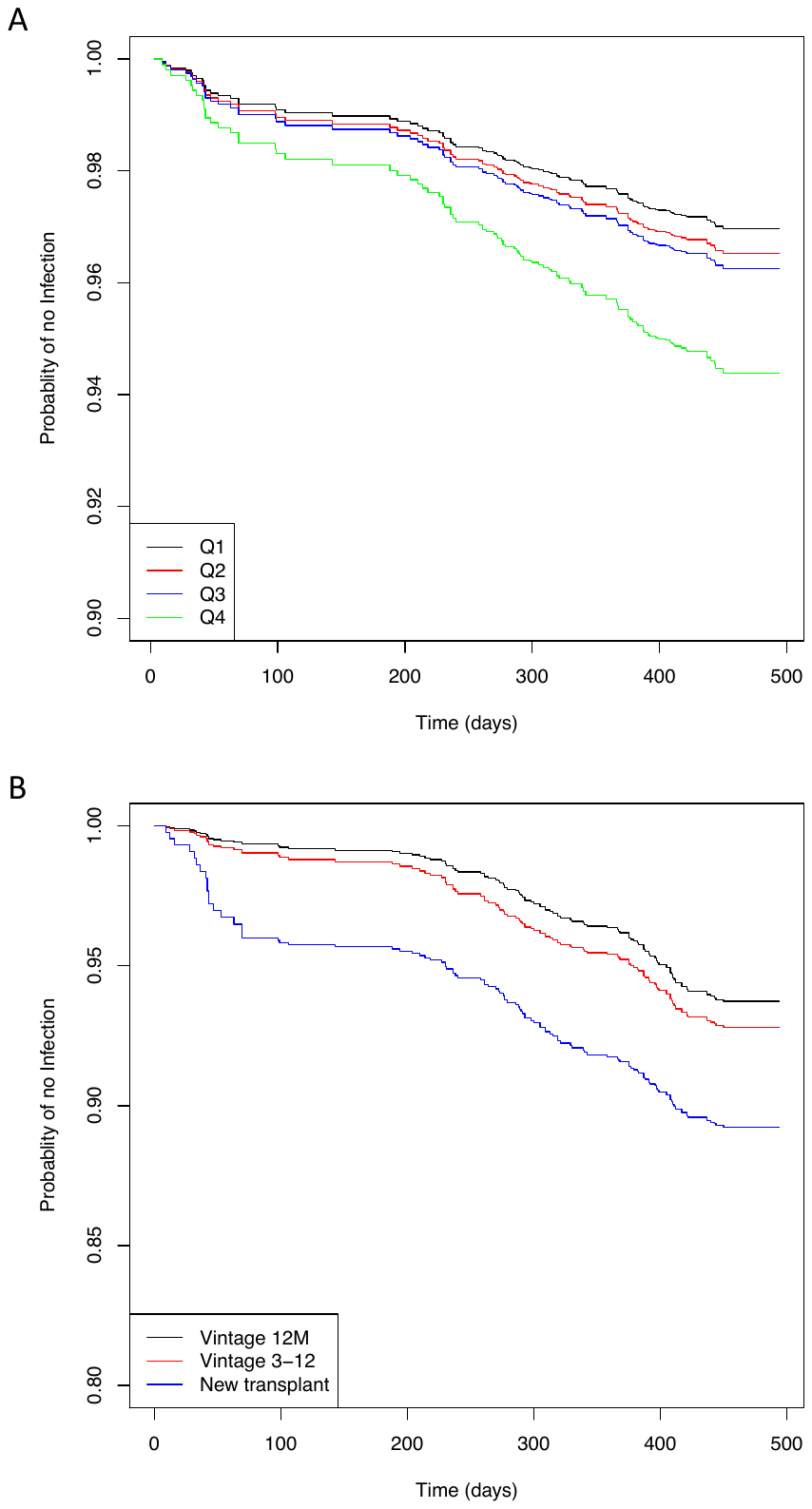

Survival curves based on the time-varying hazard function, stratified by (A) Covid-19 community burden, represented by quartiles of highest infectious risk communities and (B) transplant vintage (New transplant: 0-3 months; 3-12: 3-12 Months; >12: >12 months since transplant).

**Appendices**

**Appendix 1:** Email Survey sent to all kidney transplant recipients

Subject: UPDATE **PLEASE RESPOND ASAP** St. Michael’s Kidney Transplant Clinic: Patient Survey - COVID-19

**If you have any concerns or require assistance with completing the survey below, please only direct your calls to the following number:  416-XXX-XXXX**

**Thank you,**

**St. Michael’s Hospital Kidney Transplant Program**

**PATIENT SURVEY – COVID-19**

Dear Patient,

Recent research has shown that COVID-19 infection can be more severe in kidney transplant patients. The St. Michael’s Hospital Kidney Transplant Clinic is therefore advising all of our patients to get vaccinated unless you have a known allergy to a component of the available COVID-19 vaccines.

There is some evidence that suggests that the COVID-19 vaccines may not be as effective in transplant patients as compared to the general population. To help us understand how effective the vaccines are at protecting against COVID-19, our clinic is attempting to track COVID-19 vaccinations and infections in our patients.

To help with this effort, we have several questions below. This information will be entered into your clinical file, and will be accessible only to clinic staff. Please feel free to respond via email (preferred) with your answers.  By responding via email, you are aware and understand the limits of email confidentiality and security.

***Note - Please DO NOT call the Transplant Clinic with the COVID update information requested below.***  If we do not receive an email response by next week, the clinic will be contacting you by phone to complete the survey.

Have you received a COVID-19 vaccine?:    Yes  /  No: _______________________________

If YES, please provide responses to the following:

Manufacturer (Moderna / Pfizer / AstraZeneca / Janssen): ______________________

Date First Dose received/scheduled: ________________________________________

Date Second Dose received/scheduled:  _____________________________________

Side effect(s)? __________________________________________________________

If NO, please provide responses to the following:

is it by choice or are there barriers? _____________________________

**If there are barriers to getting the vaccine, please give us a call and we will help (416-XXX-XXXX).**

Do you have any concerns about receiving the vaccine and if Yes, what are your concerns:

__________________________________________________________

Have you had a COVID-19 infection?:    Yes  /  No: _______________________

If YES:

Date of positive test: ___________________________________________

Were you hospitalized? _________________________________________

Did you need to be put on a ventilator? ____________________________

Did you require transfer to an intensive care unit (ICU)? _______________

Finally, our program does a lot of research.

Would it be okay for someone from our research team to contact you some day if there is a study that might be of interest to you? If you say yes, you will not be contacted very often.  Yes  /  No:  ______________________

Thank you for filling out this information. We are always here to support you.

St. Michael’s Hospital Kidney Transplant Program

***This email is intended only for the named recipient(s) and may be confidential. Any unauthorized use, dissemination or copying of this email is strictly prohibited. If you have received this email in error, please notify the sender immediately and destroy all copies of this email.***

**References**

1.	Austin, P.C., Lee, D.S. & Fine, J.P. Introduction to the Analysis of Survival Data in the Presence of Competing Risks. *Circulation* **133**, 601-609 (2016).

2.	Team, R.C. R: A language and environment for statistical computing. (R Foundation for Statistical Computing, Vienna, Austria.

2020).
